# Safety Profile of Sinopharm COVID-19 Vaccine and Breakthrough Infections in Pakistan

**DOI:** 10.1101/2022.01.18.22268965

**Authors:** Wajiha Rizwan, Ahmad Uzair Qureshi, Muhammad Nasir Rana, Mubeen Nazar Duggal, Muhammad Sohaib, Masood Sadiq

## Abstract

**Background:** To determine the safety profile of Sinopharm COVID-19 vaccine and identify breakthrough infections.

**Method:** The study design was analytical cross sectional. An online questionnaire was filled by 1033 respondentsbetween 16th and 22nd April 2021. Adults who had received both doses of Sinopharm COVID-19 vaccine more than a week ago or only a single dose with serious side effect were included in the study. The frequency and severity of vaccination related side effects were assessed and breakthrough infection identified.

**Results:** The mean age of participants was 36.7 ± 12.91(18 – 92) years. Ninety one percent of participants (n=946) were health care professionals. One fifth (n=225/1033, 21.8%) had suffered from COVID-19 infection prior to vaccination, confirmed using the nasal RT-PCR test. None of the participants reported serious (grade III) or life threatening (grade IV) adverse reactions after either of the two doses. The most common side effects after the first dose were pain at injection site (20.3%), fatigue (20.3%), headache (13.9%), myalgia (12.5%) and fever (9.3%) whereas after the second dose were fatigue (16.8%), pain at injection site (15.8%), myalgia (14%) and fever (6.7%). The side effects were more common in participants who had previous history of COVID-19 infection. Of 225 previously infected participants, 97(43.1%) (p value=0.020) and 90 (40%) (p value=0.001) participants had side effects after 1^st^ and 2^nd^ dose respectively. 16 participants (1.55%) developed PCR positive COVID-19 infection two weeks after the second dose while 3(0.29%) participants had a re-infection. There was one case of probable severe COVID-19 infection, 2 weeks after the second dose and recovered completely with treatment.

**Conclusion:** Our study shows that Sinopharm COVID-19 vaccine is generally safe with no serious side effects. The side effects were however, more common in inviduals who already had COVID-19 infection. The COVID-19 breakthrough infection and reinfection could occur after the vaccination.

## Introduction

Severe acute respiratory syndrome coronavirus 2 (SARS-Cov-2) also named as Coronavirus disease 2019(COVID-19) is a rapidly transmissible disease that affected millions around the globe.^1^ Although preventive measures like social distancing, using face masks and widespread lockdowns led to control over COVID-19 rapid spread to varying extent, still paradoxically it left most people without immunity against this disease and at risk of another wave of illness.^2^ The rapid development of vaccines against SARS-CoV-2, stands out as an exceptional success story in the era of historic COVID-19 pandemic.^3^ Over a dozen vaccines have got approval for emergency use in different countries, having impressive range of efficacy from 50 - 95%.^4, 5^ All of these conditionally approved vaccines till now mostly had only temporary mild to moderate adverse reactions. The most commonly reported local side effect is tenderness or pain at injection site whereas fever, fatigue or body ache are most common systemic adverse effects.^5, 6^

Although mRNA vaccines (Pfizer-BioNTech/Moderna) were the first kind to get emergency approval having efficacy of almost 95% and an excellent safety profile noted during phase III trials, they require ultra-cold storage that limits vaccine availability in low and middleincome countries. Whereas inactivated, non-replicating viral vector and protein subunit COVID-19 vaccines have slightly less efficacy but relatively permissive cold storage requirements, making their deployment process easier in less developed countries.^5^ The success of any vaccine campaign depends on public willingness to get vaccinated. Factors that influence the vaccine acceptance include complacency, confidence on effectiveness and safety of vaccine and convenience. Convenience implies the affordability, availability and easy delivery of vaccine.^7^

In Pakistan, BBIBP-CorV, also known as the Sinopharm COVID-19 vaccine, an inactivated viral vaccine; got emergency-use approval on 18^th^ January 2021 and vaccination campaign started on 2^nd^ February 2021.^5,8^ Vaccine was recently approved by World Health Organization’s (WHO) emergency use listing on 7^th^ May 2021. The Sinopharm vaccine is given as two intramuscular doses, administered at an interval of 3 weeks. Reportedly it has an efficacy of 79% against the symptomatic SARS-CoV-2 infection, 2 weeks after the second dose.^8, 9^ Pakistan Government through National Command and Operation Center decided to vaccinate frontline health care professionals in the first phase of vaccination campaign followed by elderly people. The phase III trial data regarding safety and efficacy of Sinopharm vaccine was not published at that time and due to lack of pragmatic evidence regarding its safety and efficacy there was a lot of vaccine hesitancy.^10^ We conducted this online survey to determine its safety profile and identify side effects in different groups of vaccine recipients and dose of vaccine.

## Material and Methods

A cross-sectional study (online survey) was conducted from 16^th^ April to 22^nd^ April 2021 after taking ethical permission and approval from institutional review board of The Children’s Hospital and Institute of Child Health, Lahore (Ref. No 6761, dated 06-02-21).

At the time of conduction of the study only health care workers and senior citizens were allowed to get registered vaccination for in Pakistan. We only included participants who had completed their vaccination and were ready voluntarily to participate. The study population included adults who have received both doses of Sinopharm COVID-19 vaccine more than a week ago or only one dose in case of a serious side effect after the first dose. The form requested participants to give consent for study assuring that their confidentiality will be maintained. The participants were informed that their identity will not be included and all identifiers were removed during publication. There was no conflict of interest of researchers and there was no affiliation with or involvement in any entity or organization with any financial interest.

A questionnaire was designed on Google forms in simple English language based on the literature review ^11, 12^ after discussion among the research team.The questionnaire inquired about background data of respondents, such as age, gender, and electronic mail (e-mail) address, history of hypertension, ischemic heart disease(IHD), known allergies and past COVID-19 infection.The information was gathered regarding frequency and severity of side effects after first and second dose of Sinopharm vaccine. We graded side effects as mild (does not interfere with activity), moderate (interference with activity or repeated use of non-narcotic pain reliever >24 hours), severe (requiring narcotic pain reliever or preventing daily activity) or serious (life threatening requiring emergency room visit or hospitalization) according to FDA toxicity grading scale for volunteers enrolled in preventive vaccine trials.^13^ We also gathered information if anyone had COVID-19 infection despite vaccination with details of time period related to vaccination and onset of infection. If patient developed Polymerase Chain reaction(PCR) positive COVID-19 ≥ 2weeks after second dose, it was labeled as breakthrough infection. COVID-19 infection was classified as asymptomatic, mild, moderate, severe and critical.^14^ We used pilot study (n=20) to improve clarity of expression of survey questions. The questionnaire was then reviewed by senior members of research team and Institutional review board for face validity. The questionnaire was slightly modified as per advise of the panel and the final version was included in the manuscript.

The sample size was 631, calculated through OpenEpi software after considering frequency of side effects after Sinopharm COVID-19 vaccine 39% according to phase II trail data^12^ and confidence level 99%.

The study was conducted online by sharing the final version of questionnaire on social media (mainly WhatsApp and Facebook). The non-probability convenience sampling technique was used. The respondents were free to choose or not to participate in the survey. No money incentive was given but it was explained that their participation might prompt others to get vaccinated after knowing the safety profile of vaccine. We encouraged people to participate themselves or fill form on behalf of their household members who were eligible to participate in survey but could not fill electronic form. The respondents were also requested to pass on Google form to their acquaintances or contacts. The communication between the survey participants and investigators was carried out via e-mail when necessary The responders were also allowed to edit responses after submission.

The data was analyzed using Statistical Package for the Social Sciences (SPSS) version 25. The data was first exported from the excel sheets to SPSS. All variables were categorized according to their characteristics.. Descriptive statistics of socio demographic characteristics and variables regarding vaccination were presented as mean, standard deviation and ranges. Frequency percentages were calculated for discrete variables including side effects, COVID-19 infection and reinfection rates. The data was then distributed based on gender and age group strata. Any correlation between comorbidities and the frequency and severity of side effects was calculated using the software inbuilt models. The differences in the frequencies and severity of side-effects and morbidities were compared and a chi-square test was used to analyze for any statistical differences among the group. P-value ≤ 0.05 was considered significant.

## Results

A total of 1033 participants filled the survey questionnaire from 16^th^ to 22^nd^ April 2021.The mean age of participants was 36.7 ± 12.91(range 18 – 92) years. They were divided into 3 age groups i.e. 18-39 years, 40-60 years and > 60 years (Table 1). Almost 2/3^rd^ (n=717-69.4%) of respondents were 18-39 years old and only (n=89-8.6%) were older than 60 years. The male to female ratio was 1.08:1. The majority of participants were health care professionals (n=946-91.6 %) mainly doctors (n=774-75%). Among the study population1/5^th^ (n=225-21.8%) had suffered from confirmed COVID-19 infection prior to vaccination. The 92(8.9%) participants had history of hypertension and 155(15.2%) participants had known allergies out of which 37 (3.58%) had asthma. Our 34(3.3%) participants had known IHD. There was no significant association between IHD and severity of side effects after first(p value =0.48) or second dose(p value =0.79) of vaccine.

**Table 1.** Socio-demographic characteristics (n=1033)

| **Variable** | **Frequency(n)** | **Percentage (%)** |
| --- | --- | --- |
| **Gender** |  |  |
| Female | 495 | 47.9 |
| Male | 538 | 52.1 |
| **Age Group** |  |  |
| 18-39 years | 717 | 69.4 |
| 40-60 years | 227 | 22 |
| >60 years | 89 | 8.6 |
| **Profession** |  |  |
| **Health Professionals** | **946** | **91.6** |
| Doctors | 774 | 75 |
| Paramedics | 64 | 6.2 |
| Medical students | 37 | 3.6 |
| Administrative staff | 29 | 2.8 |
| Nurses | 26 | 2.5 |
| Janitorial staff | 16 | 1.5 |
| **Non-Health Professionals** | **87** | **8.4** |
| Housewives | 28 | 2.8 |
| Businessmen | 20 | 1.9 |
| Retired personal | 18 | 1.7 |
| others | 21 | 2 |
| **Past COVID-19 infection** |  |  |
| Yes | 225 | 21.8 |
| No | 808 | 78.2 |

None of the participants had any serious (grade III) or life threatening (grade IV) adverse reactions like anaphylaxis after any of the two doses. After 1^st^ and 2^nd^ dose of vaccine 64.9% (n=669) and 69.6% (n=719) participants respectively, reported to have no side effects at all. On other hand, after 1^st^ dose 33.4% (n=345) had mild and 1.7% (n=18) moderate (grade II) side effects. After 2^nd^ dose, 28.8% (n=298) experienced mild and 1.5% (n=16) moderate side effects (Table 2). The most common side effects after the first dose were pain at injection site (20.3%, n=210), fatigue (20.3%,n=210), headache (13.9%, n=144), myalgias (12.5%, n=129) and fever (9.3%(n=96). Whereas after second dose most common side effects included fatigue (16.8%, n=174), pain at injection site (15.8%, n=163), myalgias (14%, n=145) and fever (6.7%, n=69).

**Table 2.** Reported side effects of Sinopharm COVID-19 Vaccine.

| Side effects | Frequency (n and %) |  |
| --- | --- | --- |
|  | 1<sup>st</sup> dose | 2<sup>nd</sup> dose |
| No side effects | 669 (64.8%) | 719 (69.6%) |
| Mild side effects | 346 (33.5%) | 298 (28.8%) |
| Moderate side effects | 18 (1.7%) | 16 (1.5%) |
| Severe side effects | 0 (0%) | 0 (0%) |
| Serious side effects(Life threatening) | 0 (0%) | 0 (0%) |
| **Local Symptoms** |  |  |
| Pain at Injection site | 210 (20.3%) | 163 (15.8%) |
| Rash at Injection site | 4 (0.38%) | 3 (0.3%) |
| Itching at injection site | 19 (1.8%) | 15 (1.45%) |
| Numbness in arm | 1 (0.96%) | 0 (0%) |
| **Non-specific Systemic Symptoms** |  |  |
| Fever | 96 (9.3%) | 69 (6.7%) |
| Fatigue | 210 (20.3%) | 174 (16.8%) |
| Myalgias | 129 (12.5%) | 145 (14%) |
| Joint Pain | 31 (3%) | 33 (3.2%) |
| Sweating | 14 (1.3%) | 11 (1.06%) |
| Palpitations | 18 (1.7%) | 9(0.9%) |
| Chills | 18(1.7%) | 8 (0.77%) |
| Generalized itchiness | 6 (0.58%) | 4 (0.4%) |
| **Respiratory Symptoms** |  |  |
| Flu | 44 (4.3%) | 38 (3.7%) |
| Cough | 25 (2.4%) | 51 (4.9%) |
| Difficulty in breathing | 7 (0.67%) | 4(0.4%) |
| **GIT Symptoms** |  |  |
| Nausea | 23 (2.2%) | 20 (1.9%) |
| Diarrhea | 31(3%) | 19 (1.8%) |
| Vomiting | 4 (0.38%) | 2 (0.19%) |
| Altered taste | 24 (2.3%) | 18 (1.7%) |
| Loss of appetite | 11 (1%) | 0 (0%) |
| **Neurological Symptoms** |  |  |
| Headache | 144 (13.9%) | 119 (11.5%) |
| Dizziness | 59 (5.7%) | 48 (4.6%) |
| **Miscellaneous** |  |  |
| Blurred Vision | 5 (0.5%) | 0 (0%) |
| Elevated blood pressure | 4(0.38%) | 1(0.09%) |
| Urticarial rash | 0(0%) | 1(0.09%) |
| Conjunctivitis | 0(0%) | 1(0.09%) |
| Polyuria | 0(0%) | 1(0.09%) |

The side effects were more common in participants who had previous history of COVID-19 infection. Of 225 previously infected participants, 97(43.1%) (p value=0.020) and 90 (40%) (p value=0.001) participants had side effects after 1^st^ and 2^nd^ dose respectively. (Table 3) The frequency and severity of side effects after first dose was more among young age group (18-39 years) compared to people above 60 years of age (p value=0.047). This association was not significant after the second dose (p value=0.95%).

**Table 3.** Comparison of side effects experienced by previously COVID-19 infected and non-infected participants.

| Side effects experienced | Previously infected with COVID-19(n=225) | Previously non-infected with COVID-19(n=808) | Chi-Square | P value |
| --- | --- | --- | --- | --- |
| **After 1<sup>st</sup> dose** |  |  | **7.829** | **0.020** |
| none | 128 | 541 |  |  |
| mild | 92 | 254 |  |  |
| moderate | 5 | 13 |  |  |
| **After 2<sup>nd</sup> dose** |  |  | **13.311** | **0.001** |
| none | 135 | 584 |  |  |
| mild | 84 | 214 |  |  |
| moderate | 6 | 10 |  |  |

Among 11 lactating mothers who got two doses of Sinopharm, only one mother reported that her baby developed mild self-resolving flu after the first dose while she also had mild side effects in form of fever and flu at that time that lasted only two days after vaccination. As only mother and baby had these self-resolving symptoms immediately after vaccine and all other family members had no symptoms, she attributed them as vaccine side effect .However, no side effects noted after second dose in both of them. The mean age of these breast-feeding children was 13.9 ±5.48 months (range 4-21months).

In our study 33 participants (3.19%) had post vaccination PCR positive COVID-19 infection within two weeks after the second dose. Whereas, 16 (1.55%) respondents developed PCR positive mild breakthrough COVID-19 infection after ≥2 weeks had elapsed after the second dose (Figure1).

**Figure I.**
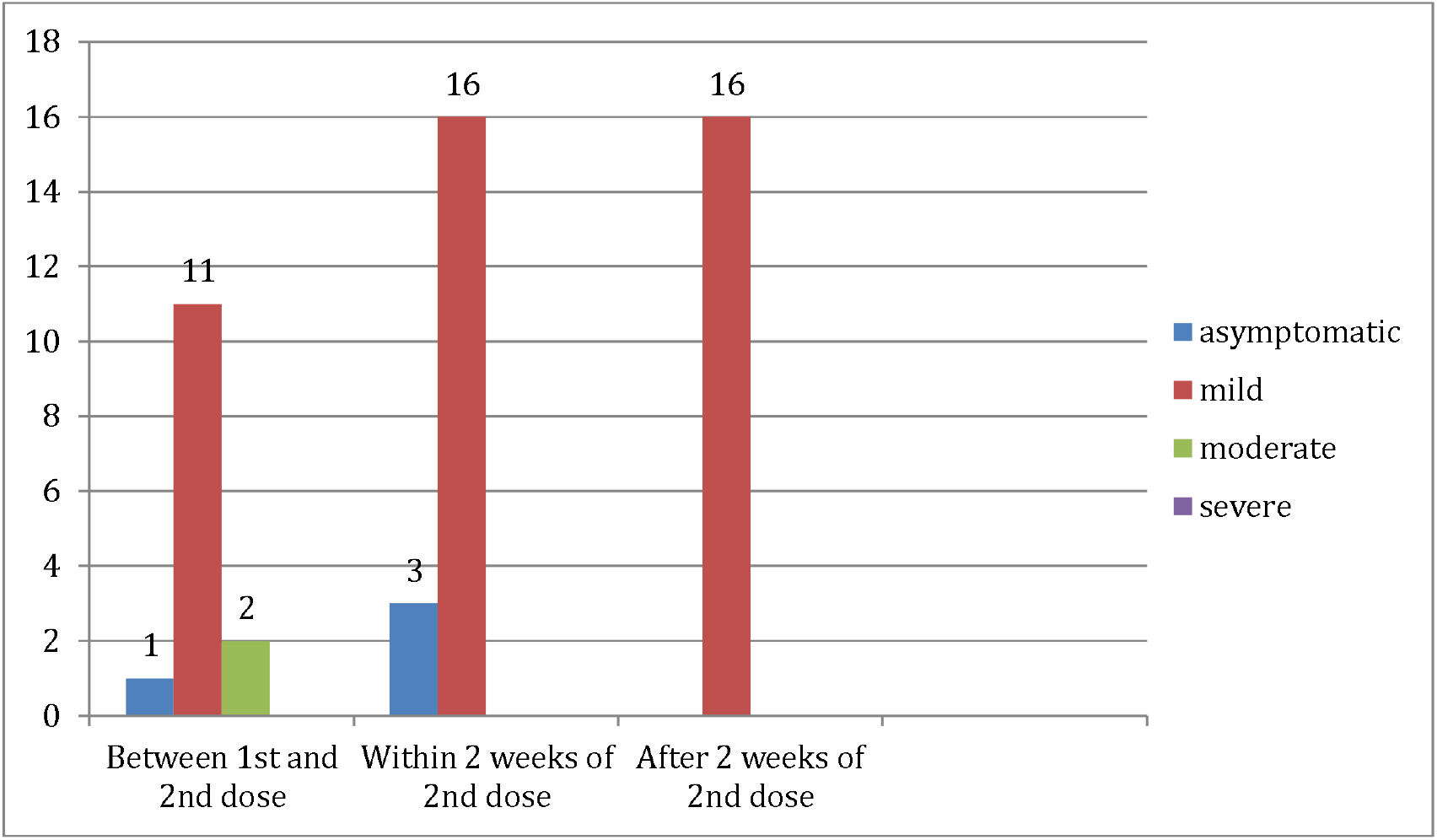
Post-Vaccination SARS-CoV-2 PCR positive infection.

Among these 16 participants, 3 also had past history of COVID-19 infection, so re-infection despite vaccination. In our study there was only 1 case of probable^15^ severe COVID-19 infection that developed infection 2 weeks after the second dose. The patients had fever, cough, shortness of breath and Oxygen saturation dropped to 84% requiring oxygen, radiological findings also suggested COVID-19 >50% lung involvement on High-resolution computed tomography of chest but as SARS-CoV-2 PCR was reported to be negative twice so we reported it as probable and not a confirmed case.

## Discussion

The mRNA Vaccines (Moderna /Pfizer-BioNTech) were amongst first FDA approved vaccines with reported efficacy of almost 95% but their distribution was mainly limited to developed and rich countries. Moreover, they require ultracold storage temperature that makes its transport and distribution difficult in in low and middle-income countries.^5^ Pakistan started its vaccination after receiving Sinopharm COVID-19 vaccine from China. It can be stored and transported at normal refrigerator temperatures. ^8^ Our study adds to the existing knowledge of its safety and is important for various resons.

Firstly, as it particularly focuses on health care workers who are more likely to report side effects completely and correctly. The 91.6 % respondents of our study were health care professionals, as the campaign started with health care professionals and during the survey time period only health care professionals and people over 60 years of age were offered Sinopharm vaccine by the government.

Secondly, the females comprised 47.6% of our study population that was much higher compared to 15.6% in phase III trial of Sinopharm conducted in the United Arab Emirates and Bahrain.^16^ The survey includes lactating mothers and their breast-fed children (albiet a small number) too as till now no Sinopharm vaccine trial included lactating mothers.^9^ It was found to be safe in this group too and we hope this positive safety evidence will encourage lactating mothers to get vaccinated.

Thirdly our results show that Sinopharm vaccine had no serious or life threatening side effect and only mild to moderate side effects were reported in 35.5% and 30.3 % participants after 1^st^ and 2^nd^ dose respectively among all age groups and gender. Phase III trials have reported similar safety profile although with some differences.^16^ The most common side effects reported after both doses were mainly pain at injection site and fatigue, whereas in phase III trial data of Sinopharm vaccine, these were mainly pain at injection site, followed by headache. The side effects were slightly more common after 1^st^ dose compared to second which is contrary to a study on side effects of Pfizer-BioNTech vaccine.^17^ Interestingly the elderly people had less side effects as compared to younger people.

Another importat aspect of our study was that the previously COVID-19 infected people had higher frequency of side effects compared to non-infected participants and this has also been observed in previous studies .^17, 18^ The reason might be that previous exposure to viral antigen leads to a robust immune response on second exposure in the form of vaccine, as also supported by observation that seropositive people developed higher antibody titers after the 1^st^ vaccine dose compared to previously non-infected individuals.^18^

The 16 participants reported to have breakthrough COVID-19 infection (i.e. PCR positive COVID-19 infection after at least two weeks of 2^nd^ dose). Among these 3 had a reinfection and all of them were young doctors working in a hospital setting but without any comorbidity. The seroprevalence studies of health-care workers have shown that the risk of infection with SARS-CoV-2 is higher (1.38 times to twice) in this group than in the general population.^19^ The limitation is lack of facility for viral genome sequencing and comparative genome analysis was not conducted in these 3 doctors. Our data supports that everyone should get vaccinated with same recommended doses irrespective of history of past COVID-19 infection.

One of our participant reported a probable severe COVID-19 infection after 2 week of second dose. She was admitted in COVID-ICU and received in addition to oxygen for 10 days remedisvir, steroids, intravenous antibiotics and oral anti-coagulant. She recovered completely. As her PCR was negative and currently to report it as breakthrough infection positive SARS-CoV-2 PCR is essential we reported it as probable case. This is also the concern showed by NA alwan^15^ that as some patients who fulfill clinical criteria for COVID-19 but either having negative PCR or not get tested are at present not included as confirmed COVID-19 cases during surveillance so our surveillance system might be underestimating the burden of COVID-19 pandemic and even missing reporting of breakthrough infections.

Even though vaccines have good efficacy but breakthrough cases are being reported. Although majority of these are non-serious infection but very few deaths have also been reported among people fully vaccinated ^21^with Food and Drug Administration(FDA) approved vaccines. Yet, COVID-19 cases and deaths that are being prevented among vaccinated person have far exceeded the breakthrough cases. ^21^

There are few limitations of our study. Firstly, the accuracy of information cannot be fully guaranteed due to self-administered online questionnaire though the recipiants were mainly health care workers, a vast majority being doctors, we performed logic check and contacted the participant to revise any data that was non-logical. We also contacted all participants who had breakthrough infection for confirmation. Secondly, the frequency of side effects might have been over estimated as we are not sure if all adverse reaction are attributable to vaccine. Thirdly, we might have underestimated the COVID-19 breakthrough infection after they filled form as we included participants who had full vaccination ≥ 1 week ago. Fourthly, the survey was done in a healthcare setting as when we conducted this online study only health care professional and senior citizens were allowed to get vaccinated. This might be reason that majority participants were doctors, so our sample might not be true representative of general population.

## Conclusions

In conclusion, Sinopharm COVID-19 vaccineis generally safe with no severe side effects. Our data also showed that the previously COVID-19 infected individuals had an increased frequency of vaccination side effects. It demonstrated that breakthrough infection and even reinfection could occur after the vaccination.

## Data Availability

All the data is available and can be provided on demand.

